# Durability and Cross-Reactivity of Immune Responses Induced by an AS03-Adjuvanted Plant-Based Recombinant Virus-Like Particle Vaccine for COVID-19

**DOI:** 10.1101/2021.08.04.21261507

**Authors:** Philipe Gobeil, Stéphane Pillet, Iohann Boulay, Nathalie Charland, Aurélien Lorin, Matthew P. Cheng, Donald C. Vinh, Philippe Boutet, Robbert Van Der Most, François Roman, Maria Angeles Ceregido, Nathalie Landry, Marc-André D’Aoust, Brian J. Ward

## Abstract

As the SARS-COV-2 pandemic evolves, what is expected of vaccines extends beyond efficacy to include consideration of both durability and variant cross-reactivity. This report expands on previously reported immunogenicity results from a Phase 1 trial of an AS03-adjuvanted, plant-based coronavirus-like particle (CoVLP) displaying the spike (S) glycoprotein of the ancestral SARS-CoV-2 virus in healthy adults 18-49 years of age (NCT04450004). When humoral and cellular responses against the ancestral strain were evaluated 6 months post-second dose (D201), 100% of vaccinated individuals retained binding antibodies, and ∼95% retained neutralizing antibodies; interferon gamma (IFN-γ) and interleukin 4 (IL-4) responses directed against the ancestral S protein were also still detectable in ∼94% and ∼92% of vaccinees respectively. Variant-specific, cross-reactive neutralizing antibody (NAb) levels were assessed at D42 and D201 using both live wild-type and pseudovirion assays (Alpha, Beta, Gamma) or the wild-type assay alone (Delta, Omicron). In the wild-type assay, broad cross-reactivity was detected against all variants at D42 (100% Alpha and Delta, 94% Beta and Gamma, 74% Omicron). At D201, cross-reactive antibodies were detectable in almost all participants against Alpha, Gamma and Delta variants (94%) and the Beta variant (83%) and in a smaller proportion against Omicron (44%). Results were similar in the pseudovirion assay (D42, 100% cross-reactivity to Alpha and Gamma variants, 95% to Beta variant, D201, 94% for Alpha, Beta and Gamma variants). These data suggest that two doses of 3.75 µg CoVLP+AS03 elicit a durable and cross-reactive response that persists for at least 6 months post-vaccination.

## Introduction

Coronavirus disease 2019 (COVID-19), the disease caused by severe acute respiratory syndrome coronavirus 2 (SARS-CoV-2), has spread rapidly across the globe, infecting more than 492 million people and including 6.2 million deaths as of March, 2022 ^1^. The disease typically affects the upper and lower respiratory tracts, where it can cause severe clinical features including dyspnea, hypoxemia, tachypnea, lung edema, and acute respiratory failure ^2^. Multiple cellular and molecular mediators of immune responses, inflammation, and coagulation appear to be involved in the pathogenesis ^3^.

Vaccination against SARS-CoV-2 remains an effective strategy for preventing viral transmission and reducing disease severity, hospitalizations, and deaths ^4^. COVID-19 vaccines based on at least six different platforms have been explored, with more than 300 SARS-CoV-2 vaccine candidates in pre-clinical and clinical developmental, 37 of which have received emergency use approval in at least one country ^5,6^.

Reported vaccine efficacies in large field trials early in the pandemic when the ancestral strain dominated ranged from ∼50–95% ^7,8^ and were highly correlated with serum levels of both binding antibodies and neutralizing antibodies (NAb) against the ancestral strain ^9-12^ which were proved to be highly predictive of neutralization of variants of concern ^13^. The inverse correlation between antibody titers and viral load has also been observed in animal challenge models in which vaccine-generated antibody titers are associated with restricted viral loads and reduced lung inflammation ^14,15^. Furthermore, the transfer of antibodies from convalescent to naïve animals ^16^ or between humans in a clinical setting ^17,18 preprint^ can result in a decline in viral loads, reduced symptoms and lower mortality. More recently, evidence has been steadily accumulating that cell-mediated immune effectors also contribute to both short-term protection and the establishment of long-lasting immunity ^19-21^.

Although no correlate of immunity has been widely accepted, the persistence of circulating antibodies and cell-mediated immune responses after natural infection and/or vaccination may be indicative of durable protection. Following infection, NAb titers decline gradually after the initial peak but remain detectable in most individuals for up to 16 months ^22-25^. Studies of vaccine-induced humoral immunity suggest a similar pattern with an estimated half-life between 50-60 days depending on the antibody parameter assessed and the model used ^26,27^. Although antibodies with broad neutralizing activity can be induced naturally in some individuals, the antibodies generated in most people for most viral infections tend to be highly specific ^28^. In contrast to the relatively short-lived and specific humoral response, cellular memory responses (ie: antigen-specific CD4 T cells) are present in most individuals after SARS-CoV-2 infection for at least 8 months ^22,29^. After the 2002-2003, SARS-COV outbreak, interferon-γ (IFN-γ) ELISpot responses were readily detectable in most subjects for at least 6 years post-infection ^30^. By their nature, T cell responses tend to be highly cross-reactive ^31,32^.

As many countries move from a primary pandemic response to a mixed pandemic/endemic response, both the durability and breadth of protection induced by vaccines become increasingly relevant. This work expands on the previously described Phase 1 study evaluating the plant-based CoVLP+AS03 vaccine candidate ^33^ by reporting the persistence of cell-mediated immune (CMI) responses to ancestral strain S protein antigens up to 6 months post-vaccination as well as the durability and cross-reactivity of vaccine-induced NAbs against Alpha, Beta, Gamma, Delta and Omicron variants ^34^.

## Results

Participant demographics are detailed in Table 1. A panel of human convalescent sera/plasma (HCS) from patients recovering from mild, moderate, or severe COVID-19 infection (sampled 27 to 105 days post symptom onset) is included for comparison. This report builds on the reported short-term (up to D42) Phase 1 antibody and CMI responses against the ancestral strain ^33^. No deaths, study vaccine-related SAEs, AESIs, or AEs leading to withdrawal were reported up to Day 386 of the study. One SAE, an adenocarcinoma of colon, reported ∼3.5 months post-vaccination occurred in a subject in the 15 µg unadjuvanted group; this event was assessed as not related to study vaccine by the Investigator and the Sponsor. Thus, no late-onset events of concern were identified with the CoVLP vaccine. No safety signal of concern has been detected in the study through Day 386. The safety results from this study support continued investigation of CoVLP as a vaccine candidate for the prevention of SARS-CoV-2 infection. Herein we report the persistence of humoral response against the ancestral strain to six months (D201) and one year (D386) after vaccination. The persistence of the cellular response was measured after six months. We also report the short-term (D42) binding cross-reactivity to SARS-CoV1, MERS, and common cold coronaviruses as well as both short-(D42) and long-term (D201) NAb cross-reactivity to Alpha, Beta, Gamma, Delta, and Omicron variants.

**Table 1:** Summary demographics and baseline characteristics of the trial subgroup of participants who received 3.75 µg CoVLP adjuvanted with AS03 (NCT04450004) and patients convalescing from COVID-19.

| Comparisons | Healthy individuals | Convalescent individuals |  |  |
| --- | --- | --- | --- | --- |
|  | 3.75 µg CoVLP with AS03 adjuvant | Mild | Moderate | Severe |
| Subjects, n | 20 | 16 | 8 | 11 |
| Sex, n (%) |  |  |  |  |
| Male | 5 (25.0) | 10 (62.5) | 2 (25.0) | 8 (72.7) |
| Female | 15 (75.0) | 5 (31.3) | 6 (75.0) | 3 (27.3) |
| Information non-available |  | 1 (6.2) |  |  |
| Race, n (%) |  |  |  |  |
| White | 20 (100.0) | 7 (43.8) | 6 (75.0) | 5 (45.5) |
| Black or African American | 0 (0.0) | 1 (6.3) | 0 (0.0) | 5 (45.5) |
| Asian | 0 (0.0) | 4 (25.0) | 1 (12.5) | 0 (0.0) |
| Ethnicity, n (%) |  |  |  |  |
| Hispanic/Latinx | 0 (0.0) | 2 (12.5) | 1 (12.5) | 1 (9.1) |
| Age, years |  |  |  |  |
| Mean ± SD | 34.7 ± 9.1 | 42.7 ± 13.6 | 37.8 ± 13.0 | 51.9 ± 16.0 |
| Median (range) | 36 (19–49) | 39 (20–66) | 40.5 (19–58) | 50.0 (28–82) |

CoVLP, plant-produced virus-like particle; SD, standard deviation.

### Durability of Humoral Responses

To evaluate the durability of the humoral response against the ancestral strain, the anti-spike immunoglobulin G (IgG) enzyme-linked immunosorbent assays (ELISA), pseudovirion neutralization assays (PNA), and live-virus microneutralization assays (MNA) were used, as previously described ^33^.

Spike-binding IgG to the ancestral strain were detected in all participants at D42 and D201 (19/19 and 18/18, respectively; Figure 1a). Similarly, for both the PNA and the MNA assays, D201 NAbs were present in almost all participants (17/18; 94%) and were not significantly different from proportions at D42 (18/18; 100%; p>0.9999 for both assays; Figures 1b and 1c).

**Fig. 1.**
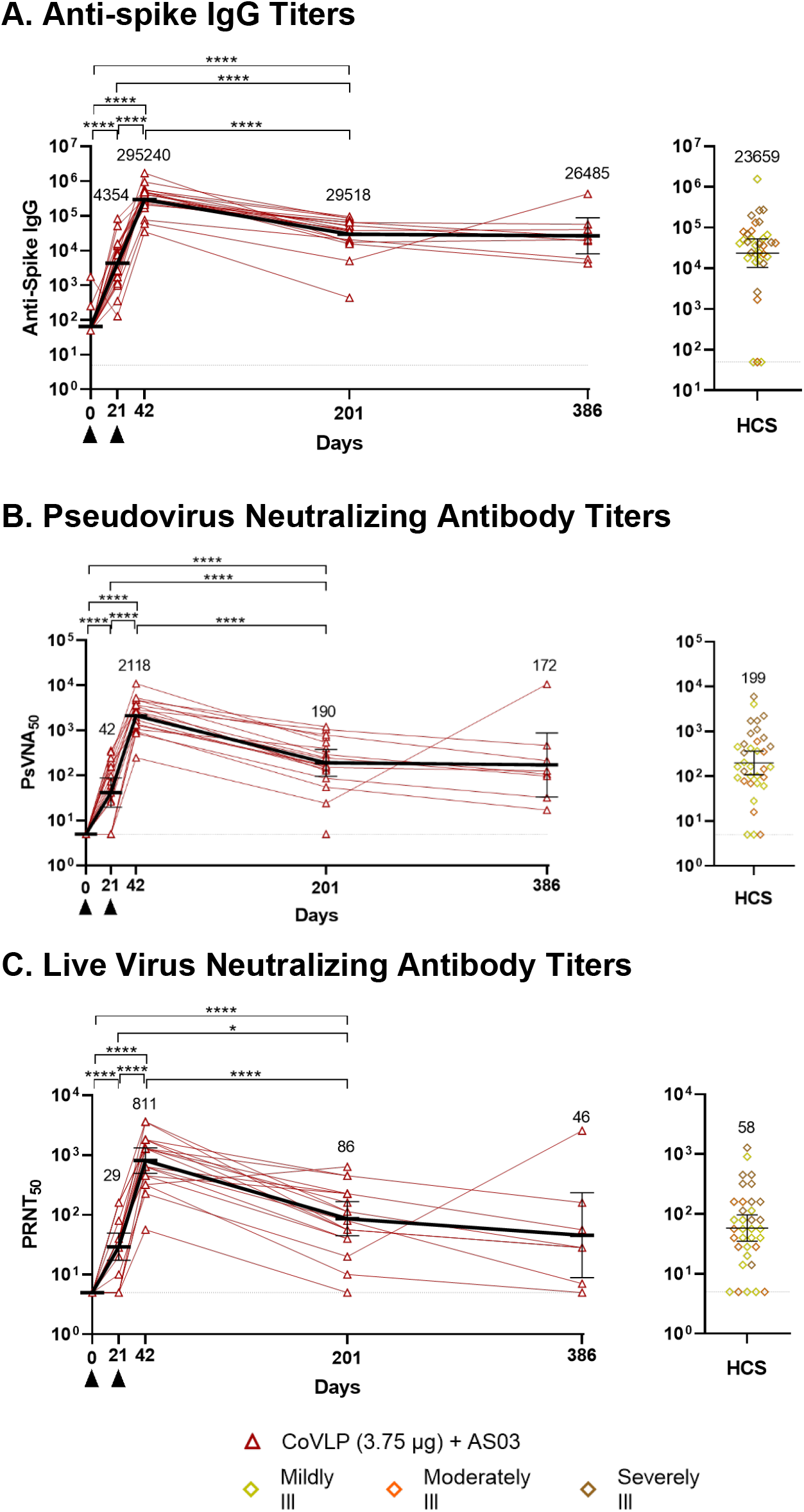
Durability of antibody responses. Antibodies in serum collected at Day 0, 21, 42, 201 and 386 post first immunization from subjects vaccinated with 3.75 µg CoVLP adjuvanted with AS03 were measured by ELISA (Immunoglobulin G against S protein, panel A). Neutralizing antibodies were measured using ancestral (Wuhan) strain derived vesicular stomatitis virus pseudovirus (panel B) or live virus (panel C) -based assays. Values from convalescent sera or plasma (HCS) collected at least 14 days after a positive diagnosis of COVID-19 (RT-PCR) from individuals whose illness was classified as mild, moderate, or severe/critical (n=35) are shown in the right-hand panels. Individual data are indicated (red lines) along with geometric means (horizontal black lines and numerical values). For panel A, vaccinated subjects at D0 and 21 (n=20); at D42 (n=19); at D201 (n=18); at D386 (n=8) and HCS (n= 34) were included. For panel B, vaccinated subjects at D0 and D21 (n=20); at D42 (n=18); at D201 (n=18); at D386 (n=8) and HCS (n= 35) were included. For panel C, vaccinated subjects at D0 and D21 (n=20); at D42 (n=19); at D201 (n=18); at D386 (n=8) and HCS (n= 35) were included. Error bars indicate 95% CI. Black triangles indicate immunization. Significant differences between timepoints are indicated by asterisk(s) (*p<0.05; ****p<0.0001; One-way analysis of variance using a mixed-effect model on log-transformed data GraphPad Prism, v9.0). Due to the limited sample size at D386, that timepoint was not included in the statistical analysis.

The D201 anti-spike IgG geometric mean titer (GMT) (29,518; 95% confidence interval [CI]: 17,938-48,574) was significantly lower (p<0.0001) than the D42 GMT (295,240; 95% CI: 137,96–631,790; Figure 1a) although remaining significantly (p<0.0001) higher than the response at D21 (4,354, 95% CI: 2,070-9,159), and comparable to the HCS (23,659; 95% CI: 10,579-52,909). In order to allow wider context and interpretation of the binding antibody data, World Health Organization (WHO) pooled plasma 20/136, composed of mixed convalescent plasma ^35^, was included as a reference standard. Based on these results, a normalization factor was applied to transform the GMT values to binding antibody units per milliliter (BAU/mL). The post-vaccination sera had a GMT of 5,350 BAU/mL on D42 and 535 BAU/mL at D201. The measured GMT value for HCS was 428.8 BAU/mL.

On D201, the GMT values for the PNA and the MNA assays were comparable to HCS despite a decline from peak D42 values (Figures 1b-c). On D201, 17 of 18 (94.4%) individuals retained PNA titers and the (GMT 190; 95% CI: 96.03-377.0) was significantly lower (p<0.0001) than at D42 (2,118; 95% CI: 1,229-3,652) although remaining significantly (p<0.0001) higher than the response at D21 (42; 95% CI: 27-64) and comparable to the HCS GMT (199; 95% CI: 109-364; Figure 1b). Similarly, the D201 GMT in the MNA (86.4; 95% CI: 56.4-132.4) was significantly lower (p<0.0001) than the D42 value (811; 95% CI: 496-1,327) but similar to that of HCS (58.3; 95% CI: 35.1-96.8; Figure 1c). Using the WHO mixed convalescent plasma reference standard (20/136), a normalization value was applied to the PNA results to obtain GMT values of 1,131 International Units per milliliter (IU/mL) at D42 and 101 IU/mL at D201 post-vaccination. The corresponding value for HCS was 106 IU/mL. Similarly, a normalization value was applied to the MNA data to obtain GMT values of 896 IU/mL at D42 and 95.5 IU/mL at D201. The corresponding value for the HCS was 64.1 IU/mL.

D386 serum samples from 8 individuals were tested for Spike-binding IgG antibodies by ELISA and neutralizing antibodies against the ancestral strain Spike protein by PNA and MNA. Due to the limited sample size, no statistical analysis was conducted relative to this timepoint (Figures 1a-c). ELISA analysis of binding antibodies from D386 serum samples (Figure 1a) revealed seropositivity in all 8 samples (100%) and yielded a GMT of 26,485 (95% CI: 11,883-59,033). PNA analysis (Figure 1b) revealed seropositivity in all 8 samples (100%) and a GMT of 172 (95% CI: 79.2-372). MNA analysis (Figure 1c) revealed seropositivity in 7 of 8 samples (87.5%) and a GMT of 45.6 (22.6-91.7).

The half-lives (*t*_½_) of the vaccine-generated anti-spike IgG binding and NAbs were calculated using the exponential-decay model. The *t*_½_ values for antibodies in all three assays were comparable, with overlapping 95% CIs: 55.26 days for anti-spike IgG (n=18; 95% CI: 44.67-65.85), 56.44 (n=17; 95% CI: 44.08-68.80) for the PNA and 59.23 days (n=16; 95% CI: 39.83-78.63) for the MNA.

Overall, these data show that two doses of CoVLP+AS03 elicited binding antibodies and NAbs that remained detectable 1 year after the second dose. Antibody titers at both D201 and D386 were comparable to those seen in patients recovering from natural COVID-19 infection.

### Cross-Reactivity to SARS-CoV-1, MERS, and Common Cold Coronaviruses

Figure 2 shows the reactivity for the spike proteins of SARS-CoV-2, SARS-CoV-1, and Middle East Respiratory Syndrome (MERS) of serum antibodies 21 days after the second immunization with CoVLP+AS03 (D42) compared with HCS measured using the fluorescence-based multiplex VaxArray platform from InDevR (Colorado, USA).

**Fig. 2.**
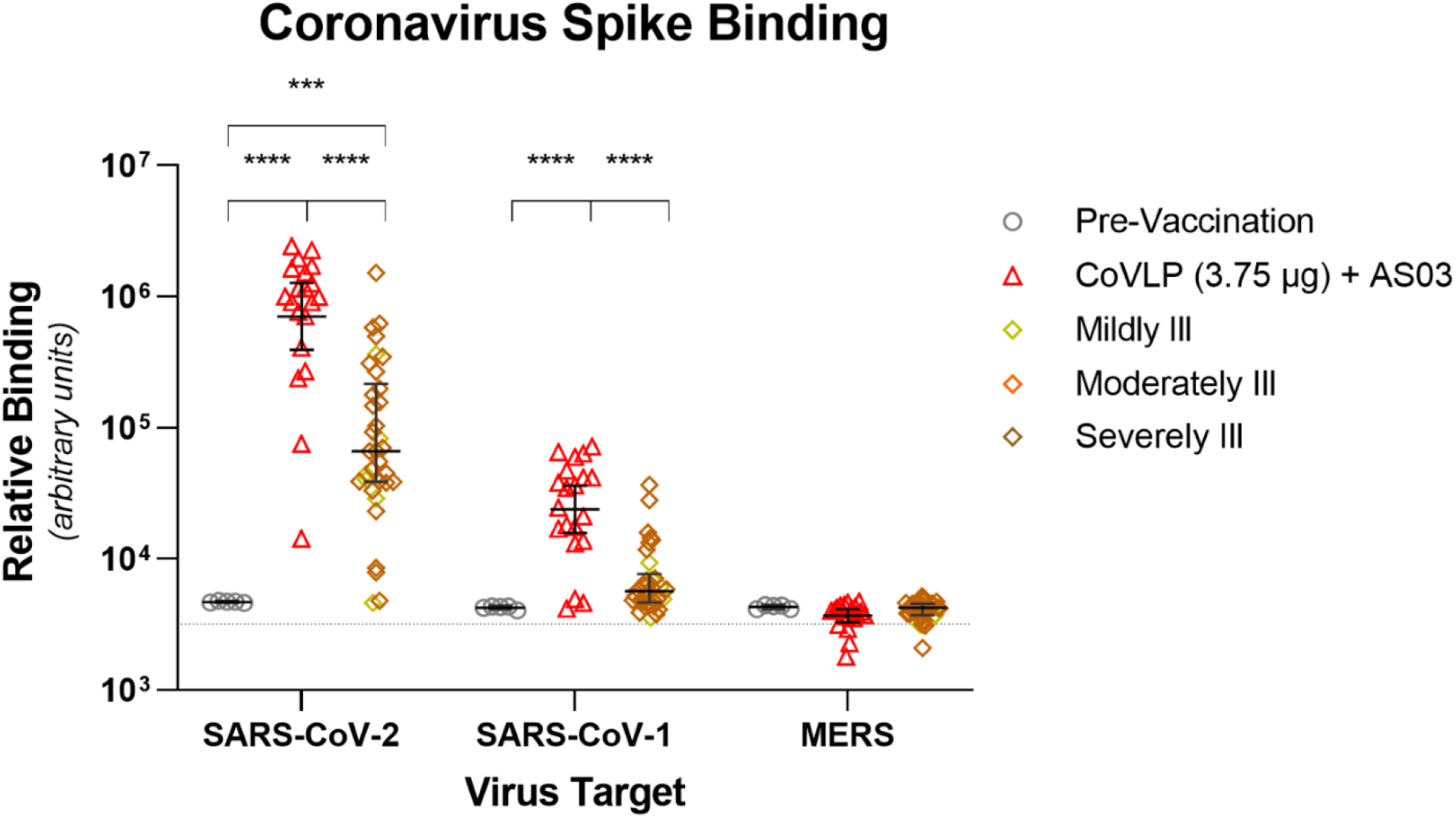
Binding antibody cross reactivity to betacoronaviruses. Binding of serum antibodies from pre-vaccinated subjects (n=5) and from D42 of subjects vaccinated with 3.75 µg CoVLP adjuvanted with AS03 (n=20) to protein S from SARS-CoV-2, SARS-CoV-1, and MERS (geometric mean and 95% CI) were quantified using the VaxArray platform from InDevR, Inc. Convalescent sera or plasma collected at least 14 days after a positive diagnosis of COVID-19 (RT-PCR) from individuals whose illness was classified as mild, moderate, or severe/critical (n=35) were analyzed concurrently. Dotted line indicates mean background control values. Significant differences between sera are indicated by asterisks (***p<0.001; ****p<0.0001; One-way analysis of variance on log-transformed data, GraphPad Prism, v9.0).

As expected, pre-vaccination sera were not reactive to the spike proteins of SARS-CoV-2, SARS-CoV-1, or MERS. Sera from subjects vaccinated with CoVLP+AS03 and patients recovering from COVID-19 were highly reactive to the SARS-CoV-2 spike protein in this assay. Antibody binding for vaccinated individuals were approximately one order of magnitude higher than for individuals in the HCS group (*p*<0.0001).

Although the binding of vaccinated and HCS sera to the SARS-CoV-1 spike protein was lower than to the SARS-CoV-2 spike protein, sera from vaccinated individuals still had significantly higher binding to the SARS-CoV-1 spike protein than HCS (*p*<0.0001).

Neither vaccination nor infection with SARS-CoV-2 induced significant cross-reactive antibodies to the MERS spike protein (Figure 2) or spike proteins from common cold coronaviruses (Supplemental Figure 1).

### Cross-Reactivity to Variant of Concern

Cross-reactive NAb induced by CoVLP+AS03 against Alpha, Beta, Delta, Gamma and Omicron variants were assessed at D42 and D201.

Results in the PNA for the ancestral strain as well as the Alpha, Beta, and Gamma variants revealed a similar pattern of cross-reactivity (Figure 3a). At D42, all vaccinated individuals had readily detectable cross-reactivity to Alpha (GMT 1,544; 95% CI: 908-2,626) and Gamma variants (GMT 555; 95% CI: 344-895). Cross-reactivity to the Beta variant was observed in 18 of 19 participants (94.7%: GMT 273; 95% CI: 140-535). At D201, 17 of 18 (94.4%) individuals retained NAb titers to all of the variants tested using the PNA: Alpha (GMT 177; 95% CI: 91.6-343), Beta (GMT 65.7; 95% CI: 38.0-114), and Gamma (GMT 121; 95% CI: 66.3-220).

**Fig. 3.**
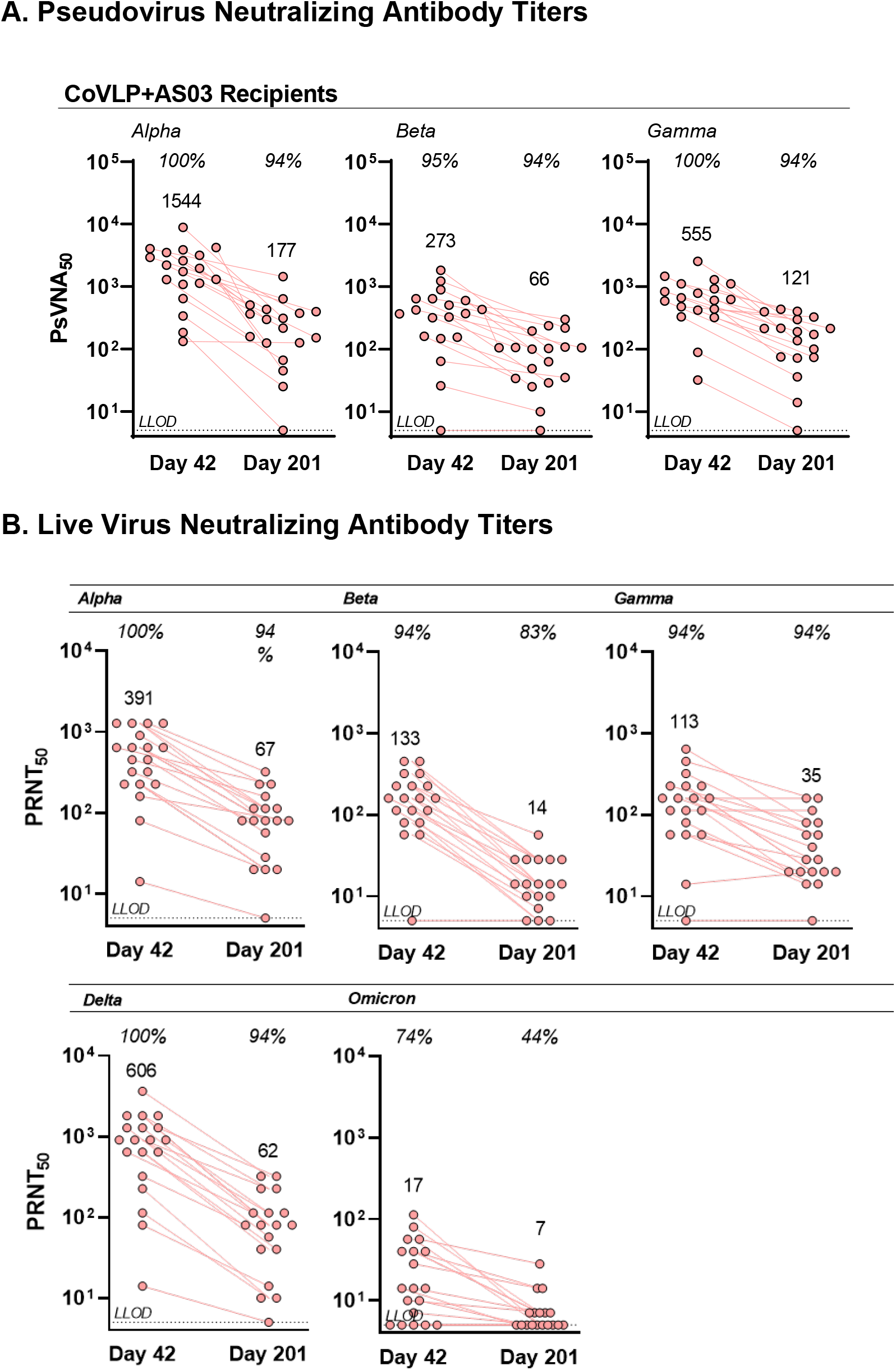
Neutralizing antibody cross-reactivity to SARS-CoV-2 variants. The cross-reactivity of NAbs in subjects vaccinated with 3.75 µg CoVLP adjuvanted with AS03 (n=19 at D42, n=18 at D201) were quantified by (A) VSV pseudovirion neutralization assay against the Alpha, Beta, or Gamma variants or by (B) live virus neutralization assay against the Alpha, Beta, Gamma, Delta and Omicron variants. Individual values are represented; geometric means are indicated above each series of datapoints. Percentage seropositivity relative to the variant being tested are shown at the top of each dataset.

Using a live virus neutralization assay (Figure 3b), NAb were readily detected at D42 in 19 of 19 (100%) participants to Alpha and Delta variants, and 18 of 19 (94.7%) participants to the Beta and Gamma variants. Cross-reactivity to the antigenically distinct Omicron variant was observed in 14 of 19 (73.7%) participants. The D42 GMTs to Alpha, Beta, Gamma, Delta and Omicron variants were 811 (95% CI: 497-1327), 391 (95% CI: 228-672), 133 (95% CI: 82.0-217), 113 (95% CI: 65.2-196), 606 (95% CI: 320-1147), and 17.3 (95% CI: 10.4-28.8) respectively. At D201, persistent reactivity was observed in 17 of 18 (94.4%) participants against Alpha (GMT 67.3; 95%CI: 39.8-114), Gamma (GMT 35.0; 95% CI: 21.9-55.7) and Delta variants (GMT 62.3; 95% CI: 33.8-115). Cross-reactivity to the Beta variant at D201 was observed in 15 of 18 (83.3%: GMT 14.1; 95% CI: 10.0-20.1) individuals and to the Omicron variant in 8 of 18 individuals (44.4%: GMT 6.8; 95% CI: 5.3-8.7).

Taken together, these results show that 2 doses of the CoVLP+AS03 vaccine given 3 weeks apart induced a NAb response to ancestral strain that persisted in most individuals for up to 386 days post-vaccination. The 2 doses of Medicago vaccine candidate also induced cross-reactive antibodies against Alpha, Gamma and Delta variants that persisted in the large majority (∼95%) of participants at D201. The induction and persistence of cross-reactive NAb were generally lower to the antigenically distinct Beta and Omicron variants although responses against Omicron were still measured in 44% of the participants 6 months after the first immunization.

### Durability of Cellular Immune Responses

The CMI response and associated T_h_1/T_h_2 balance was evaluated by expression of IFN-γ (T_h_1) and IL-4 (T_h_2) by PBMC upon *ex-vivo* re-stimulation using a SARS-CoV-2 spike-derived peptide pool (Wuhan strain; Figure 4). At D201, almost all participants had a readily detectable IFN-γ response (17/18; 94%), comparable to the proportion of IFN-γ responders at D42 (19/19; 100%). Similarly, at D201, the large majority of participants had detectable IL-4 response (12/13; 92%), again comparable to the proportion of responders at D42 (19/19; 100%). Like the humoral response, the magnitude of the cellular response was reduced on D201 relative to D42. The D201 median IFN-γ spot-forming units per million PBMCs (SFU/10^6^) response of 202.5 (95% CI: 62–433) was significantly reduced (p<0.05) relative to the D42 value of 628 SFU/10^6^ (95% CI: 403–862). Similarly, at D201, the IL-4 median SFU/10^6^ value of 46 (95% CI: 8– 151) had also fallen significantly (p<0.05) compared to the D42 median SFU/10^6^ value of 445 (95% CI: 339–680). Despite the reduced magnitude of response at D201, ongoing spike-specific IFN-γ and IL-4 cellular responses in the majority of participants suggested that two doses of CoVLP+AS03 can induce a durable CMI response.

**Fig. 4.**
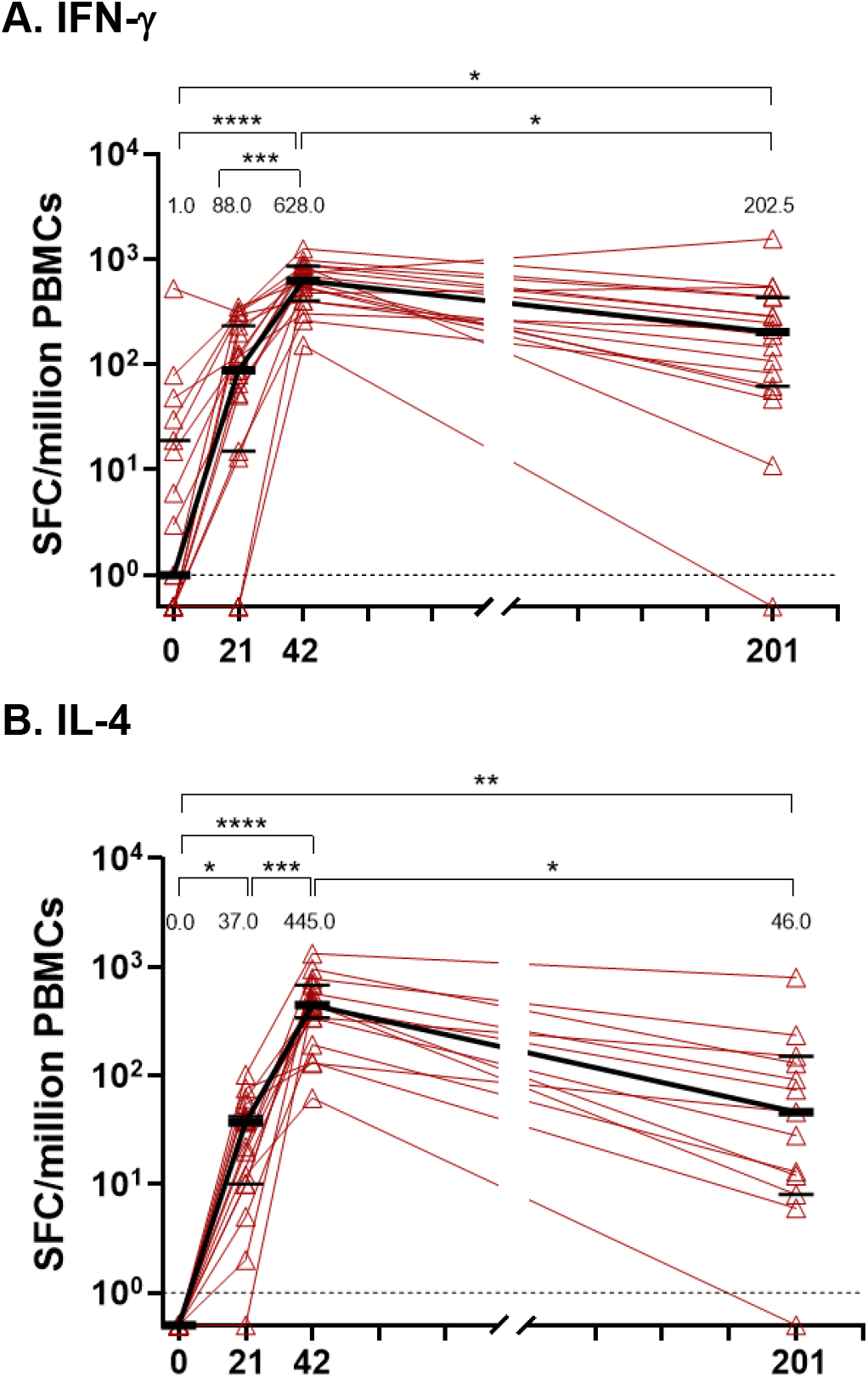
Durability of cellular immune responses. Antigen-specific IFN-γ and IL-4 responses were quantified by ELISpot. Frequencies of spike-specific cells producing IFN-γ and IL-4 per million PBMCs at baseline (D0) and 21 days after one immunization (D21), or two immunizations (D42), and D201 post-immunization with 3.75 µg CoVLP adjuvanted with AS03 were measured after *ex vivo* restimulation with a peptide pool consisting of S protein-derived 15-mer peptides overlapping by 11 amino acids. For IFN-γ, vaccinated subjects (n=19) at D0, 21, 42 and at D201 (n=18) were included in the analysis. For IL-4, vaccinated subjects at D0 (n=14), D21 (n=17), D42 (n=19) and at D201 (n=13) were included in the analysis. Individual values are indicated by red lines; medians are indicated by black lines and numerical values. Error bars indicate 95% CI. Significant differences between timepoints are indicated by asterisk(s) (*p<0.05; **p<0.01; ***p<0.001; ****p<0.0001; Friedman test followed by Dunn’s comparisons test, GraphPad Prism, v9.0).

## Discussion

Early vaccine trials conducted during the initial waves of the SARS-CoV-2 pandemic demonstrated efficacies between 70-95% ^36,37^. More recently however, both clinical trials and real-world evidence have demonstrated that, despite remaining effective in the prevention of severe COVID-19 manifestations, overall vaccine efficacy is lower, likely driven by both the evolution of immune-evasive variants and waning immunity with time^38,39^.

In Medicago’s recent Phase 3 trial conducted during a period dominated by Gamma and Delta variants, CoVLP+AS03 demonstrated an overall efficacy of 71.0% against any symptomatic disease and of 78.1% against moderate-to-severe disease (86% in those seronegative at baseline) ^40^. When sequence information was available, variant-specific efficacy of CoVLP+AS03 was observed to be 75.3% and 88.6% for the Delta and Gamma variants respectively and 100% for smaller number of Alpha, Lambda and Mu variants ^40^. The viral loads in the nasal passages of the breakthrough cases in this study were >100-fold lower than those in the placebo cases suggesting that the vaccine had significant virologic impact even if it did not completely protect against mild illness. The work presented here broadens our understanding of the possible role that CoVLP+AS03 may be able to play in the dynamic environment of emerging and evolving variants and transient vaccine-induced immunity.

Overall, the humoral immune response induced by CoVLP+AS03 was demonstrated to be robust, durable, and cross-reactive. As previously reported ^33^, both serum S-binding and neutralizing antibody levels at 21 days after the second dose of CoVLP+AS03 were ≥10-fold higher that those seen in patients recovering from natural disease. Although there are obvious concerns about the comparability of humoral responses between studies that use different assays and report fold-differences using study-specific panels of sera, the peak binding antibody titers elicited by CoVLP+AS03 are among the highest reported for any vaccine when expressed as standardized BAUs: 5350 BAU/mL at D42 and 535 BAU/mL at D201 ^41^. In this context, it is noteworthy that when Feng et al used the same WHO pooled reference standard (ie: 20/136) ^42^ to correlate binding antibody titers with efficacy against the ancestral strain ^43^, they reported that a titer of 264 (95% CI: 108-806) BAU/mL was predictive of 80% vaccine efficacy against symptomatic COVID-19. In a similar study, Goldblatt and colleagues ^44^ estimated the threshold for protecting against symptomatic COVID-19 was 154 (95% CI: 52-559) BAU/mL.

While there are advantages and limitations to each approach, whether the antibody responses are reported as fold-difference versus convalescent serum ^45^, as standardized BAU values, or as the proportion of participants with detectable levels (ie: binding assay 100% and NAb 94% at D201), the humoral response to CoVLP+AS03 was both robust and durable.

Based on the GMTs at D42 and D201, the calculated *t*_½_ of the anti-spike IgG binding and NAbs of 55-60 days were comparable to antibody decay results reported after either natural disease ^24,46^ or administration of other SARS-CoV-2 vaccines ^26^. The limited number of samples available from D386 sampling precludes a fair quantitative comparison relative to earlier timepoints. Nevertheless, it is reassuring to observe that 87.5-100% of the serum tested remained positive (depending on the assay used) and that the rate of antibody decay appears to slow substantially between D201 and D386. This is consistent with similar biphasic antibody decay curves that have been reported for several other vaccines with a rapid decline in most subjects for the first 3-6 months followed by a much slower decline thereafter ^47-49^.

In the context of the evolving nature of the ongoing pandemic, it is reassuring that the humoral response induced by CoVLP+AS03 was highly cross-reactive. Although there was no cross-reactivity to the endemic human coronaviruses or MERS, as has been reported for natural SARS-CoV-2 infection ^50^, there were readily-detectable responses to the original SARS-CoV (100% of subjects) and substantial cross-reactivity against a broad array of variants both at D42 (range 74-100% with detectable responses) and at D201 (range 44 – 94%). As might be expected given their mutational differences, cross-reactivity was higher against the Alpha and Delta variants, was reduced but still substantial against the Beta and Gamma and lowest against the Omicron variant. While the live virus and pseudovirion neutralizing antibody outcomes assessed in this study were consistent for the ancestral strain and the Alpha, Delta and Gamma variants, it is notable that cross-reactivity to the Beta variant was more nuanced. Anti-Beta neutralization at D201 was higher in the pseudovirion assay (94% with detectable titers at D201) relative to the live virus neutralization assay (83%). The proportion of participants with cross-reactive neutralizing antibodies to Omicron at D42 was reduced compared to other variants (74%) but was in the same range as has been reported for other approved vaccines. Although the proportion of participants retaining cross-reactivity fell to 44% at D201, this observation was similar to reported results for other available vaccines ^51,52^. While no Omicron-specific efficacy data are available yet from the Phase 3 trial of CoVLP+AS03, the vaccine performed well against a range of variants: 75% (Delta), 89% (Gamma) and 100% (Alpha, Lambda, Mu) in sequence-positive, symptomatic cases ^40^.

The addition of AS03 to protein-based vaccine has been demonstrated to increase cross-reactivity and duration of the immune response ^53^. However, the mechanism(s) by which the combination of CoVLP and AS03 induces this robust, durable, and cross-reactive humoral response remains a subject of investigation. Based on the recently demonstrated variant-specific efficacy of CoVLP+AS03 ^40^ and the cross-reactivity described herein, there may be potential benefit to incorporating CoVLP+AS03 into the current public health strategy^54^.

While a great deal of attention has been paid to humoral responses induced by SARS-CoV-2 vaccines, the role of T cells has been relatively under-studied. Nonetheless, it is clear that Th1-type responses play an important role in recovery from acute viral infections including highly pathogenic coronaviruses ^20^ and early T cell responses after vaccination can greatly influence both the magnitude and quality of the short-term response as well as the induction of long-term memory ^22,29,55^. Although the experience with SARS-CoV-2 is limited to ∼24 months, T cell responses are readily detected for at least 12 months in many patients recovering from either SARS-CoV ^56^ or SARS-CoV-2 ^57^. Indeed, persistent T cell responses were found for up to a decade following SARS-CoV infection in 2003-2004 ^58^. Given the intrinsic cross-reactivity of T cell responses and the on-going challenge of the SARS-CoV-2 variants, the ability of a vaccine to elicit both antibody and T cell responses has become increasingly relevant ^59,60^. In the current study, CoVLP+AS03 vaccination rapidly induced both Th1 and Th2 responses that persisted for at least 6 months after the second dose in all but one participant.

While the IL-4 response observed in this study is quite prominent compared to responses reported following other vaccines ^7^, the balance between Th1 (IFN-γ) and Th2 (IL-4) responses favored a Th1-type response after each dose (Figure 4) and the Th1/Th2 ratio increased over time to D201, allaying concerns about possible vaccine-associated enhanced disease (VAED) ^61,62^. Although Th-2 skewing was initially considered a potential risk for SARS-CoV-2 vaccines and assessment of T_h_1/T_h_2 polarization was considered essential during early COVID-19 vaccine development ^63,64^, to our knowledge no suggestion of VAED nor, *at fortiori*, association between inappropriate Th2 response and VAED have been observed with any SARS-CoV-2 vaccine ^65^. In the case of CoVLP+AS03 specifically, there has been no suggestion of VAED in either a macaque challenge study ^15^ or in the clinical trials conducted to date despite the strong induction of both Th1- and Th2-type cytokine responses ^33,40,66^. In fact, the IL-4 response elicited by the combination of CoVLP and AS03 may play an important role in the strength of the antibody responses observed, possibly by supporting follicular T-cell involvement and germinal center development ^67-70^. Indeed, AS03 administered with other antigens has been shown to promote broad Th1- and Th2-type cytokine responses that contribute to both the strength and the durability of humoral responses ^71-73^.

Although characterization of the cellular response presented herein are limited to ELISpot data, a more detailed analysis of the T cell response to CoVLP+AS03 vaccination is underway and will be reported separately.

Vaccines that can induce cross-reactive and durable responses are likely to play an increasingly important role as the SARS-CoV-2 pandemic evolves towards endemicity. The data presented herein suggest that CoVLP+AS03 can induce such a response and provide mechanistic support for the efficacy recently demonstrated by this vaccine against a wide range of variants of concern ^40^.

## Methods

### CoVLP vaccine candidate and adjuvant

The CoVLP vaccine candidate has previously been described in detail ^33^. Briefly, full-length spike protein from SARS-CoV-2 (strain hCoV-19/USA/CA2/2020) incorporating the modifications R667G, R668S, R670S, K971P, and V972P is expressed in *Nicotiana benthamiana* by transient transfection, resulting in spontaneous trimer formation, VLP assembly and budding. The purified CoVLP is mixed with AS03 immediately prior to injection. The AS03 adjuvant is an oil-in-water emulsion containing DL-α-tocopherol (11.69 mg/dose) and squalene (10.86 mg/dose) and was supplied by GlaxoSmithKline.

### Study design

The Phase 1 study design investigating tolerability and immunogenicity of CoVLP with and without adjuvants was previously described ^33^. Ethical approval was provided by the Advarra Institutional Review Board as well as the Health Products and Food Branch of Health Canada and the study was carried out in accordance with the Declaration of Helsinki and the principles of Good Clinical Practices. Participants were recruited from existing databases of volunteers, and written informed consent was obtained from all study participants before any study procedure. Participants were offered modest compensation for their participation in this study (time off work and displacement costs).

### SARS-CoV-2 spike protein ELISA

This ELISA measured binding to SARS-CoV-2 S protein in its prefusion configuration (SARS-Cov2/Wuhan/2019, Immune Technology Corp.: amino acids 1-1208 with the furin site removed and no transmembrane region) as previously described ^33^.

### SARS-CoV-2 pseudovirus neutralization assay (PNA)

Pseudovirion neutralizing antibody analysis was performed using a genetically modified Vesicular Stomatitis Virus (VSV) engineered to express the SARS-CoV-2 S glycoprotein (NXL137-1 in POG2 containing 2019-nCOV Wuhan-Hu-1; Genebank: MN908947) from which the last nineteen amino acids of the cytoplasmic tail were removed (rVSVΔG-Luc-Spike ΔCT) (Nexelis, Quebec, Canada) as previously described ^33^.

Cross-reactivity to variants was tested using modified pseudovirion expressing SARS-CoV-2 S glycoprotein from Alpha (Nexelis lot #: NL2102M-N; del69-70, del144, N501Y, A570D, D614G, P681H, T716I, S982A, D1118H, plus Δ19aa C-terminal for the PP processing), Beta (Nexelis lot #: NL2103K-N; L18F, D80A, D215G, del242-244, R246I, K417N, N501Y, E484K, D614G, A701V, plus Δ19aa C-terminal for the PP processing), and Gamma (Nexelis lot #: NL-2102O-N; L18F, T20N, P26S, D138Y, R190S, K417T, E484K, N501Y, D614G, H655Y, T1027I, V1176F, plus Δ19aa C-terminal for the PP processing) variants.

### SARS-CoV-2 microneutralization CPE-based assay (MNA)

Neutralizing antibody analysis was performed using a cell-based cytopathic effect assay (VisMederi, Sienna, Italy) based on ancestral SARS-CoV-2 virus (2019 nCOV ITALY/INMI1, provided by EVAg; Genebank: MT066156) as previously described ^33^. For cross-reactivity against variants, the assay was conducted with Alpha (swab isolate 14484; mutations: N501Y, A570D, D614G, P678H, T716I, S982A, T572I, S735L, D69/70, D144Y), Beta (hCoV-19/Netherlands/NoordHolland_10159/2021), Gamma (human isolate PG_253 Clade Nexstrain 20J/501Y.V3; Mutations: L18F, T20N, P26S, D138Y, R190T, K417T, E484K, N501Y, D614G, H655Y), Delta (sab isolate 31944, mutations: G142D, E156-158del, R158G, L452R, T478K, D614G, P681R, R582Q, D950N, V1061V), and Omicron (VMR_SARSCOV2_Omicron_C1, BA.1, Mutations: A67V, H69del, T95I, G142D, V143-145del, L212I, K417N, N440K, G446S, S477N, E484A, Q493R, G496S, Q498R, N501Y, Y505H, T547K, D614G, H655Y, N679K, P681H, N764K, D796Y, N856K, Q954H, N969K) variants.

### Standardization of antibody titers with the WHO 20/136 pooled sera

As previously described ^66^, WHO International Standard anti-SARS-CoV-2 immunoglobulin (human; NIBSC code: 20/136) was included in antibody binding and neutralization assays for the purpose of facilitating comparison of results with other studies. This standard material is pooled plasma from eleven individuals who recovered from SARS-CoV-2 infection with very high NAb responses ^42^.

For the ELISA, a reference titer of 55,175 was observed; hence a normalization factor of 55.18 was used to allow expression of the ELISA results in binding antibody units per milliliter (BAU/mL).

For the PNA assay, a reference GMT value of 1872 was observed, hence a normalization factor of 1.872 was used when expressing PNA titers in international units per milliliter (IU/mL). Similarly, for the MNA assay, 20/136 generated a titer of 905.1 hence a normalization factor of 0.91 was applied to the MNA titers to allow expression in IU/mL.

### Calculation of antibody half-lives

Antibody t_½_ were calculated by exponential decay model based on values observed at D42 and D201. The mean of the individually calculated t_½_ values were reported along with 95% CI. GraphPad Prism software was used to calculate means and 95% CIs.

### Cross reactivity to SARS-CoV-1, MERS and common cold coronaviruses

Cross-reactivity to SARS, MERS and common cold coronaviruses was quantified using the VaxArray platform and the Coronavirus SeroAssay at InDevR, Inc. (Boulder, CO). Spike protein antigens representing full-length spike, receptor binding domain (RBD), and the S2 extracellular domain of SARS-CoV-2, and the spike proteins from SARS, MERS, HKU1, OC43, NL63, and 229E were printed on the microplates.

Prior to use, the microarray slides were equilibrated to room temperature for 30 minutes. All serum samples were diluted at 100-fold and a predetermined subset of 20 samples were diluted at 1000-fold in Protein Blocking Buffer (PBB) and applied to the microarray and allowed to incubate in a humidity chamber on an orbital shaker at 80 rpm for 60 minutes. After incubation, the samples were removed using an 8-channel pipette and the slides were subsequently washed by applying 50uL of Wash Buffer 1. Slides were washed for 5 minutes on an orbital shaker at 80 RPM after which the wash solution was removed. Anti-human IgG Label (VXCV-7623) was prepared by diluting the label to 1:10 in PBB after which 50uL of label mixture was added to each array.

Detection label was incubated on the slides in the humidity chamber for 30 minutes before subsequent, sequential washing in Wash Buffer 1, Wash Buffer 2, 70% Ethanol, and finally ultrapure water. Slides were dried using a compressed air pump system and imaged using the VaxArray Imaging System (VX-6000).

The slides were imaged at a 100 ms exposure time. The raw signal was converted to signal to background ratio and reported as arbitrary relative binding units.

### Interferon-γ and Interleukin-4 ELISpot

PBMC samples from study subjects were analyzed by IFN-γ or IL-4 ELISpot using a pool of 15-mer peptides with 11aa overlaps from SARS-CoV-2 S protein (USA-CA2/2020, Genbank: MN994468.1, Genscript, purity >90%). Full details of the methodology are detailed elsewhere ^33^.

### Convalescent samples

Sera/plasma from COVID-19 convalescent patients were collected from a total of 35 individuals with confirmed disease diagnosis. Time between the onset of the symptoms and sample collection varied between 27 and 105 days. Four serum samples were collected by Solomon Park (Burien, WA, USA) and 20 sera samples by Sanguine BioSciences (Sherman Oaks, CA, USA); all were from non-hospitalized individuals. Eleven plasma samples were collected from previously hospitalized patients at the McGill University Health Centre. Disease severity was ranked as mild (COVID-19 symptoms without shortness of breath), moderate (shortness of breath reported), and severe (hospitalized). These samples were analyzed in parallel with clinical study samples, using the assays described above. Demographic characteristics are presented in the Table 1.

### Statistical analysis

Humoral assays comparing data across D0, D21, D42, and D201 timepoints (Figure 1) used one-way analysis of variance using a mix-effect model of log-transformed data. Comparisons of the proportion of individuals with detectable antibodies or not were conducted using Fisher’s exact test. Analysis of antibody binding to coronavirus spike protein (Figure 2) used one-way analysis of variance on log-transformed data. Comparisons of cell-mediated immune response durability (Figure 4) across timepoints were conducted using Friedman’s test follow by Dunn’s comparisons test.

## Data Availability

Medicago Inc. is committed to providing access to anonymized data collected during the trial that underlie the results reported in this article, at the end of the clinical trial, which is currently scheduled to be 1 year after the last participant is enrolled, unless granted an extension. Medicago Inc. will collaborate with its partners (GlaxoSmithKline, Rixensart, Belgium) on such requests before disclosure. Proposals should be directed to. To gain access, data requestors will need to sign a data access agreement and access will be granted for non-commercial research purposes only.

## Acknowledgements

The authors wish to acknowledge all the volunteers who participated in the study, the site investigators and their staff who conducted the studies with a high degree of professionalism, and all the Medicago employees and contractors who were involved in the study.

## Author contributions

All authors contributed significantly to the submitted work. BJ Ward and N Landry contributed to all aspects of the clinical study from conception to completion. P Gobeil, S Pillet, I Boulay, A Lorin, N Charland, and MA D’Aoust contributed to design and execution of the study as well as analysis and presentation of the data. P. Boutet, F. Roman, R. Van Der Most, and M. de los Angeles Ceregido contributed to analysis and presentation of data. MP Cheng provided access to reagents and consulted on study design and execution. DC Vinh provided access to reagents. All authors contributed to critical review of the data and the writing of the manuscript. All Medicago authors had full access to the data. BJW made the final decision to submit the manuscript.

