## Supplemental Figure 1 for "Durability and Cross-Reactivity of Immune Responses Induced by an AS03-Adjuvanted Plant-Based Recombinant Virus-Like Particle Vaccine for COVID-19"

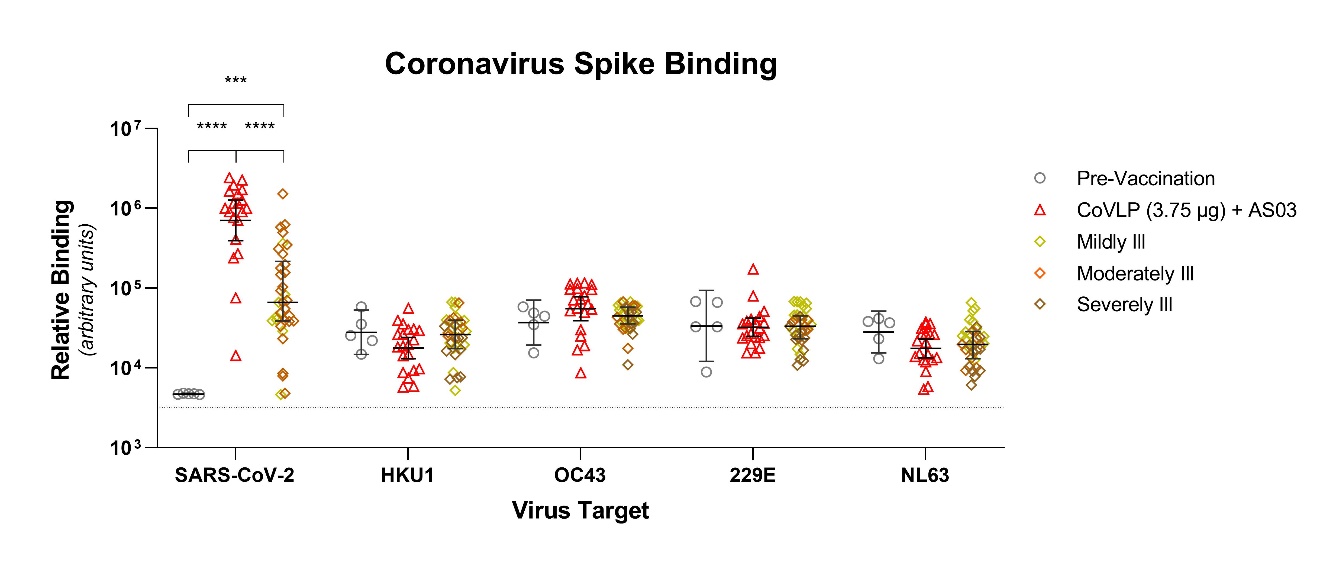

**Supplemental Figure 1:** **Neutralizing antibody cross-reactivity to common cold coronaviruses.** Binding of serum antibodies from pre-vaccinated subjects (n=5) and from D42 of subjects vaccinated (n=20) with 3.75 µg CoVLP adjuvanted with AS03 to protein S to the four common cold coronaviruses (geometric mean and 95% CI) were quantified using the VaxArray platform from InDevR, Inc. Convalescent sera or plasma collected at least 14 days after a positive diagnosis of COVID-19 (RT-pCR) from individuals whose illness was classified as mild, moderate, or severe/critical (n=35) were analyzed concurrently. Dotted line indicates mean background control values. Significant differences between sera are indicated by asterisks (***p<0.001; ****p<0.0001; One-way analysis of variance on log-transformed data. GraphPad Prism, v9.0).
